# Association Between Body Roundness Index Z-Scores and the Prevalence of Hypertension in Middle-Aged and Older Adults: Evidence from a Representative Study in China

**DOI:** 10.1101/2025.01.02.25319921

**Authors:** Liangxiu Wu, Shenshen Du, Weicheng Lai, Mengxuan Liu, Yupeng Wu, Huayang Qin, Pinyou Lu, Qimeng Wu, Qian Liu, Qiao Jie, Xin Li, Liangyan Wu

## Abstract

**Introduction:** Hypertension is a global health concern and a leading risk factor for cardiovascular diseases. Central obesity, marked by abdominal fat, is more strongly linked to hypertension than general obesity. Traditional metrics like Body Mass Index (BMI) overlook fat distribution. This study examines the association between the Body Roundness Index (BRI) Z-score, a novel central obesity measure, and hypertension prevalence in China’s middle-aged and older adults.

**Methods:** Utilizing data from the China Health and Retirement Longitudinal Study (CHARLS), we included 16,102 participants aged 45 years and above. BRI was calculated and standardized into Z-scores, with participants categorized into three groups (Q1-Q3). Logistic regression and restricted cubic spline models were used to evaluate the relationship between BRI Z-scores and hypertension prevalence, adjusting for confounders.

**Results:** Higher BRI Z-score were significantly associated with increased hypertension prevalence (OR per unit increase: 1.39; 95% CI: 1.32–1.47, p < 0.0001). Subgroup analysis revealed that individuals in the highest BRI Z-score group (Q3) had a substantially elevated hypertension risk (OR: 2.33; 95% CI: 2.07–2.64). Additionally, BRI Z-score effectively identified early blood pressure abnormalities within the pre-hypertensive range (SBP 130–140 mmHg or DBP 80–90 mmHg).

**Conclusion:** The BRI Z-score outperforms BMI in assessing hypertension risk by better capturing central obesity. Integrating it into clinical and public health strategies enables early screening and personalized interventions, especially in high-risk populations. These findings emphasize the need for novel obesity metrics to tackle the growing hypertension burden globally.

## Introduction

Hypertension (HTN) is one of the most critical global public health challenges and a major risk factor for cardiovascular diseases. This chronic condition significantly increases global morbidity and mortality while imposing substantial burdens on healthcare systems and economic development[1]. With the intensification of population aging and changes in lifestyle, the prevalence of HTN continues to rise, particularly in low- and middle-income countries where its impact is most pronounced[2]. This trend exacerbates health disparities and demands urgent global public health action. Consequently, developing and implementing effective strategies for the prevention and management of HTN has become a top priority to mitigate its far-reaching consequences.

Obesity, particularly central obesity, is a significant contributor to HTN[3]. Characterized by abnormal abdominal fat accumulation, central obesity is more strongly associated with metabolic disorders and HTN than general obesity[4]. The underlying mechanisms include insulin resistance, chronic low-grade inflammation, endothelial dysfunction, and excessive activation of the sympathetic nervous system, which are pivotal pathways in the pathogenesis of HTN[5]. Although the body mass index (BMI) is widely used as a traditional measure of obesity due to its simplicity and accessibility, its limitations have become increasingly evident. BMI cannot distinguish between lean body mass and fat mass, nor can it accurately capture fat distribution, particularly in assessing central obesity and related risks, thus reducing its utility in predicting HTN risk[6].

Given the limitations of BMI, there is an urgent need to identify more precise indicators to evaluate central obesity and its role in chronic disease development. The Body Roundness Index (BRI) is a novel measure of obesity that integrates waist circumference and height, offering a more accurate representation of central fat accumulation[7]. By converting the BRI into Z-scores, this metric achieves standardization, enhancing its comparability across populations and providing a more robust tool for investigating the relationship between central obesity and chronic diseases such as HTN[8]. This approach not only facilitates more precise assessment of HTN risk but also supports the development of personalized intervention strategies. BRI Z-scores may prove particularly beneficial for targeting high-risk subgroups with a high prevalence of central obesity.

This study uses data from the China Health and Retirement Longitudinal Study (CHARLS) to examine the relationship between BRI Z-scores and HTN prevalence in adults aged 45 and above. By exploring BRI Z-scores, it aims to provide evidence for early HTN screening and personalized interventions, crucial for managing metabolic abnormalities and reducing HTN burden, especially with an aging population. This research offers new insights into the link between obesity and chronic diseases.

## Methods

### Study Design and Population

The data for participants in this study were obtained from the CHARLS, a nationally representative survey encompassing middle-aged and elderly adults from 28 provinces, 150 counties or districts, and 450 villages across China. The dataset provides extensive information, including demographic characteristics, socioeconomic status, health conditions, blood test results, and functional assessments. Baseline data were collected in 2011 (Wave 1), followed by four follow-up surveys conducted among survivors in 2013 (Wave 2), 2015 (Wave 3), 2018 (Wave 4), and 2020 (Wave 5). To date, blood tests were performed only in 2011 and 2015.

To investigate the relationship between BRI Z-score and HTN, this study utilized data from Wave 1 and Wave 3 of CHARLS. Inclusion criteria were: (1) complete HTN diagnostic data and BRI Z-score measurements; (2) age 45 years or older with comprehensive sociodemographic information. Exclusion criteria included: (1) participants younger than 45 years or with incomplete data; (2) missing data on HTN diagnosis or BRI Z-score; (3) outliers in BRI Z-score, systolic blood pressure (SBP), or diastolic blood pressure (DBP), defined as values below the mean minus three standard deviations (SD) or above the mean plus three SD. After rigorous screening, a total of 16,102 participants were included, with the detailed process shown in Figure 1.

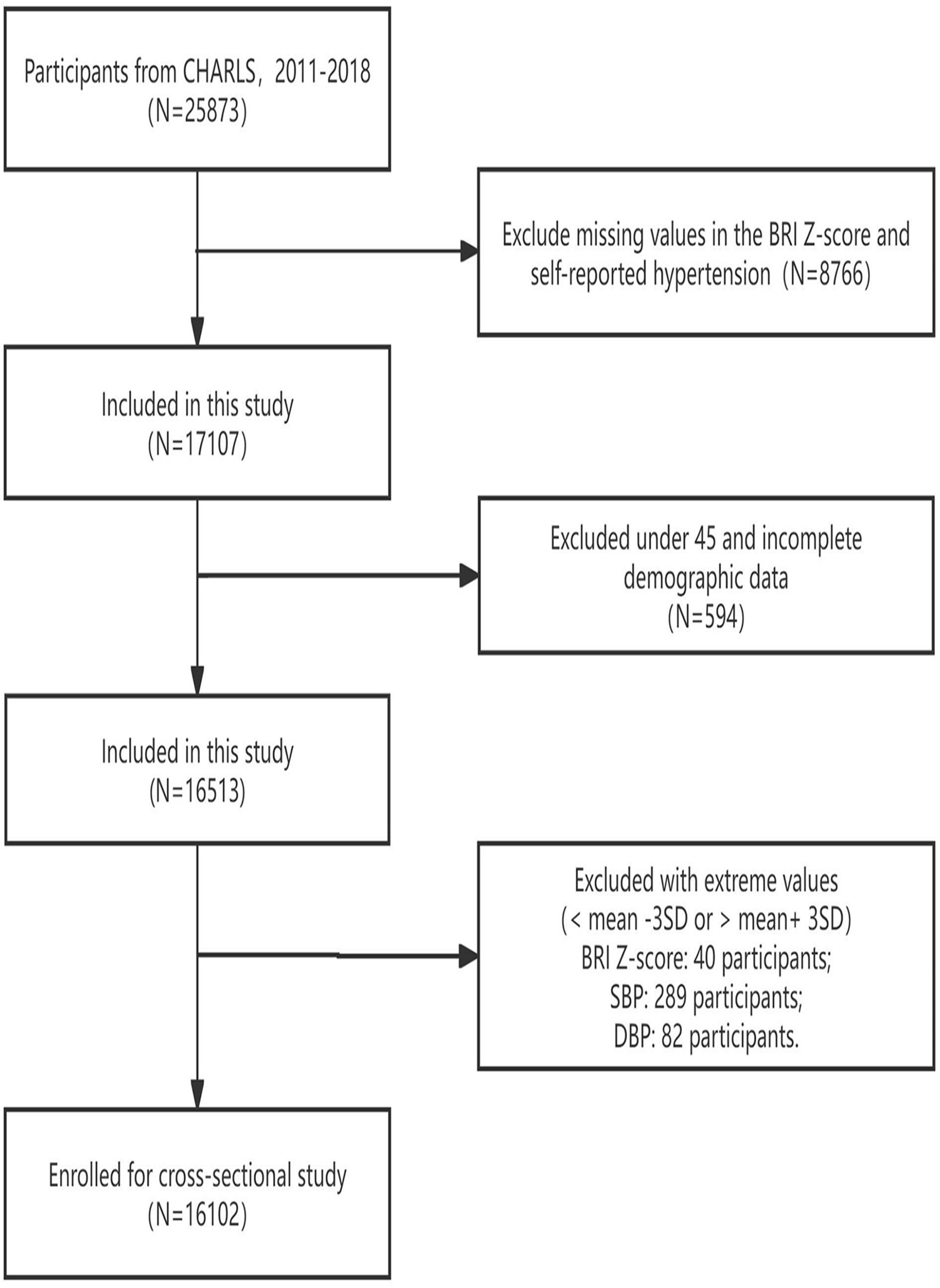

### Measurement of BRI

The BRI was calculated by[9, 10]:

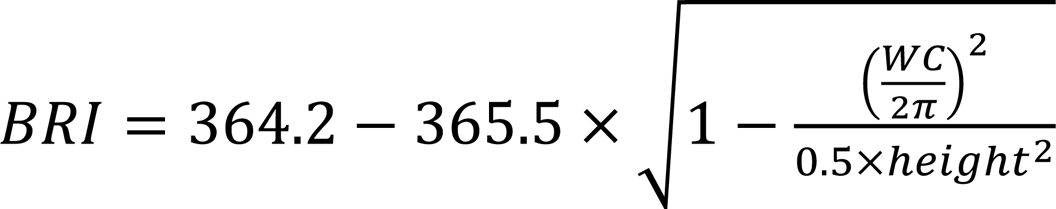

WC refers to waist circumference and both WC and Height are expressed in mmol/L.

In subsequent analysis, we considered BRI as a continuous variable, converted it into BRI Z-scores, and divided it into three groups (Q1: BRI Z-score ≤ -0.436; Q2: -0.436 < BRI Z-score ≤ 0.355; Q3: BRI Z-score > 0.355) to enhance the reliability of the analysis.

The Z-score formula is presented as follows[11]:

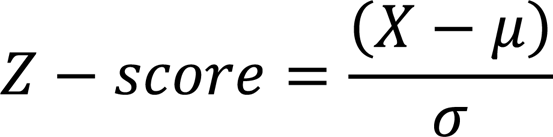

Where the data point is represented by ***X***, the mean is represented by ***μ***, and the standard deviation is represented by ***σ***.

### Ethical approval

Access to the CHARLS dataset is available on the official website (<u>charls.ccer.edu.cn/en</u>). Study protocols for biomarker collection were approved by Peking University’s Biomedical Ethics Committee (IRB00001052-11014) and the Institutional Review Board of the National School of Development, Peking University (IRB00001052-11015), with informed consent obtained from all participants.

### Data collection

Data collection for CHARLS was conducted by professionally trained personnel through questionnaires to obtain sociodemographic information. Self-reported health-related behaviors, such as smoking and drinking status, medical history (including heart disease, HTN, diabetes, stroke, and dyslipidemia), and medication used for HTN, and diabetes were recorded. Physical measurements, including weight, height, waist circumference, and blood pressure, were performed by trained professionals. Blood pressure was measured after a 5-minute rest using an Omron HEM-7200 electronic sphygmomanometer, and the average of three readings was calculated. BMI was defined as weight (kg) divided by height squared (m²).

Fasting venous blood samples were collected to measure biochemical indicators, including fasting blood glucose (FBG), glycated hemoglobin (HbA1c), serum creatinine (Scr), C-reactive protein (CRP), total cholesterol (TC), high-density lipoprotein cholesterol (HDL-C), low-density lipoprotein cholesterol (LDL-C), triglycerides (TG), serum uric acid (SUA), and blood urea nitrogen (BUN) levels.

### Covariates

We included 10 potential confounding factors that may influence the relationship between BRI Z-score and HTN. Covariates consisted of gender, age, education level, smoking status, drinking status, marital status, TG, TC, LDL-C, and HDL-C levels.

### Definition of Hypertension

According to medical diagnostic standards, participants meeting either of the following conditions were classified as hypertensive: a SBP of 140 mmHg or higher, or a DBP of 90 mmHg or higher, in the absence of antihypertensive medication[12]. In this study, HTN was defined as a binary variable. HTN cases were identified based on the following two questions: (1) whether the participant was currently managing or treating HTN through Traditional Chinese Medicine or Western medicine; and (2) whether the participant had ever been diagnosed with HTN by a clinical doctor. Both questions required affirmative responses to confirm HTN status.

### Statistical analysis

Continuous variables were expressed as means ± standard deviations, while categorical variables were presented as frequencies and percentages. Differences between groups were assessed using one-way ANOVA, the Kruskal-Wallis H test, or the chi-square test. Three logistic regression models were used to analyze the association between different BRI Z-score groups and HTN prevalence: Model 1 was unadjusted; Model 2 was adjusted for age, gender, education level, smoking status, drinking status and marital status; Model 3 further adjusted for LDL-C, HDL-C, TG, and TC based on Model 2. Results were reported as odds ratios (ORs) with 95% confidence intervals (CIs).

This study also utilized linear trend analysis and restricted cubic spline (RCS) curves to explore the nonlinear relationship between BRI Z-score and HTN prevalence. Subgroup analyses were conducted for HTN, SBP, DBP, and pulse pressure (PP), with additional stratification by age, gender, diabetes history, alcohol consumption, smoking status, dyslipidemia, and diabetes medication use. Interaction effects were evaluated using multivariable logistic regression models. Statistical analyses were performed using Empower (version 2.0) and SPSS version 27.0 (IBM SPSS, Armonk, NY, USA), with statistical significance set at p < 0.05.

## Results

### Distribution of Average BRI and Hypertension Cases Across Regions in China

To examine the regional distribution of the average BRI and hypertension prevalence across various provinces in China, Figure 2 shows regional variations in the average BRI, with Xinjiang having the highest values (dark red) and Tibet showing lower values (gray). Figure 3 reveals that provinces like Henan and Sichuan have the highest hypertension prevalence, while Tibet and some western regions report lower rates. Regions with higher BRI values tend to have higher hypertension prevalence, suggesting a potential link between central obesity and hypertension incidence.

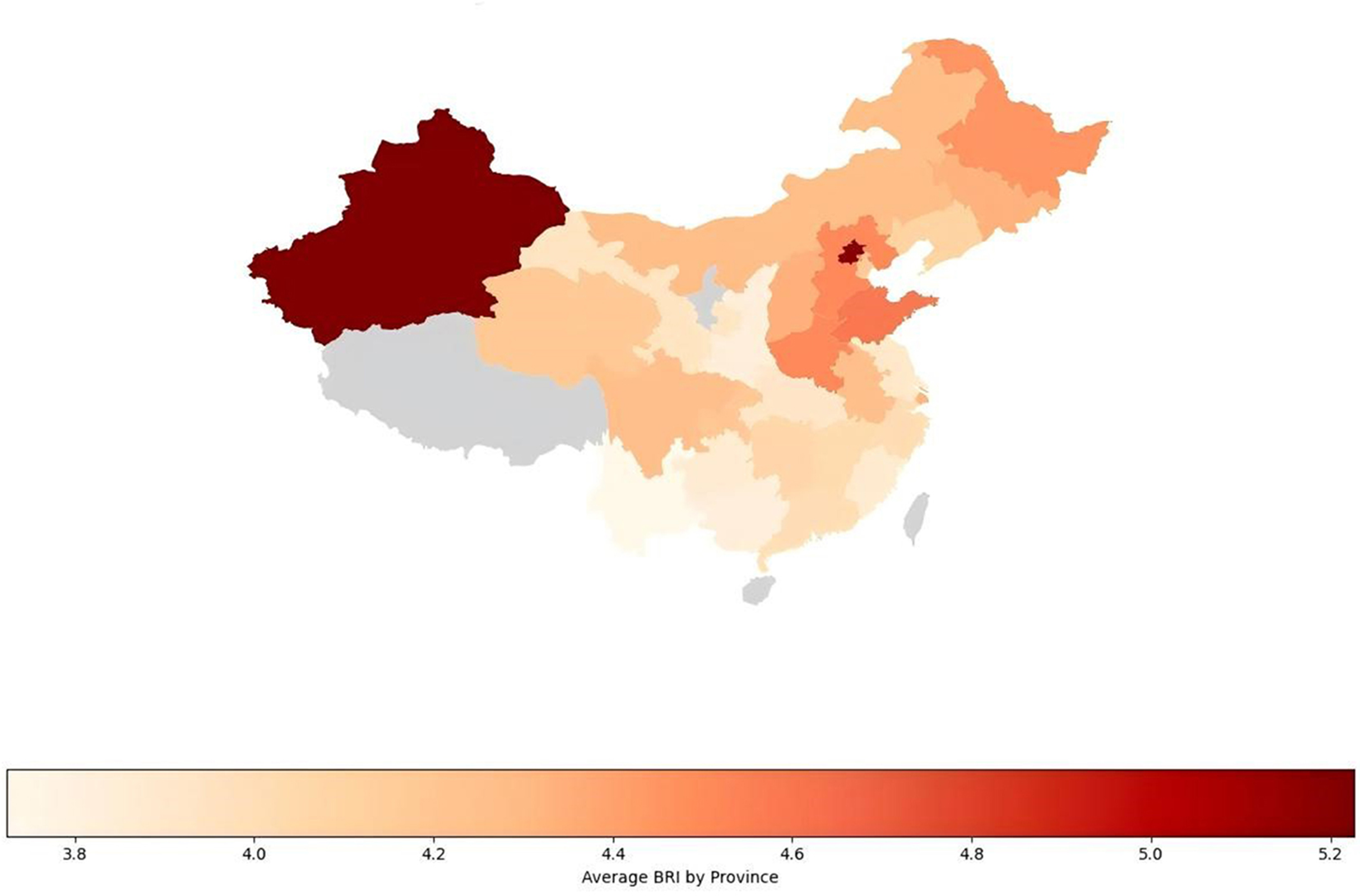

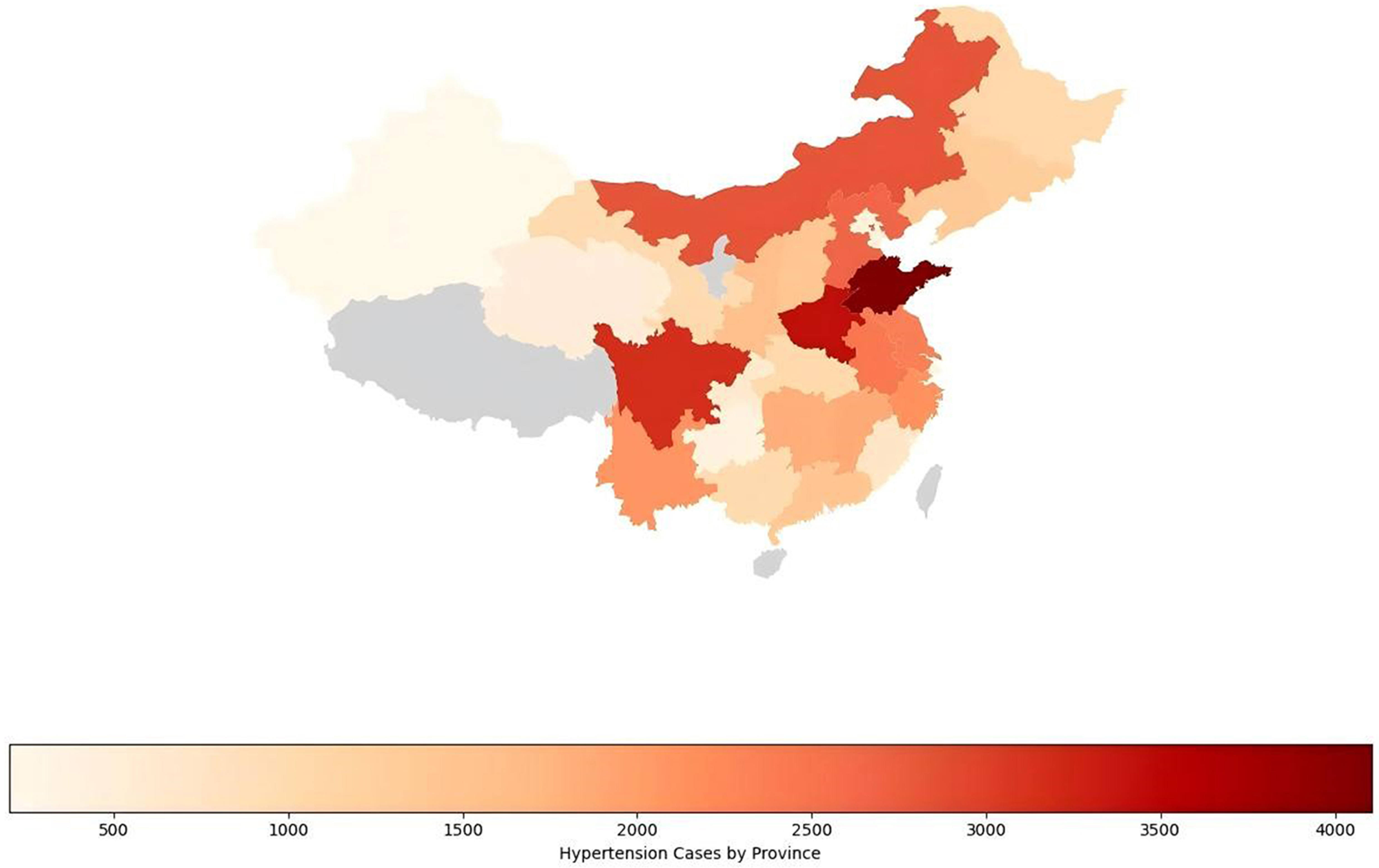

### General characteristics of the study population

A total of 16,102 participants in this study, divided into three groups according to BRI Z-score (Q1: ≤ -0.436, n=5429; Q2: -0.436 to 0.355, n=5310; Q3: > 0.355, n=5363). Results showed that the Q3 group had significantly higher levels of age, female proportion, SBP, DBP, BMI, total cholesterol, triglycerides, and LDL-C compared to the other groups, while HDL-C was highest in the Q1 group (P < 0.001). Additionally, the prevalence of HTN, diabetes, dyslipidemia, and cardiovascular disease was significantly higher in the Q3 group. In terms of health behaviors, smoking and alcohol consumption rates were higher in the Q1 group, whereas the use of antihypertensive and antidiabetic medications was more common in the Q3 group. All differences were statistically significant (P < 0.001), indicating that a higher BRI Z-score is associated with greater metabolic and disease risk (Table 1).

**Table 1.** Baseline characteristics of participants BRI Z-score in cross-sectional study.

| Variable | Total | Q1 | Q2 | Q3 | *p*-value |
| --- | --- | --- | --- | --- | --- |
|  |  | BRI Z-score≤ -0.436 | -0.436>BRI Z-score≤ 0.355 | BRI Z-score> 0.355 |  |
| Participants, sample size (N) | 16102 | 5429 | 5310 | 5363 | NA |
| Age, years | 59.34 ± 9.56 | 59.07 ± 9.60 | 58.64 ± 9.38 | 60.30 ± 9.70 | <0.001 |
| Gender (%) |  |  |  |  | <0.001 |
| Male | 7747 (48.05) | 3511 (64.67) | 2596 (48.89) | 1640 (30.58) |  |
| Female | 8355 (51.95) | 1918 (35.33) | 2714 (51.11) | 3723 (69.42) |  |
| Education level (%) |  |  |  |  | <0.001 |
| Below high school or vocational school | 14316 (88.91) | 4833 (89.02) | 4641 (87.40) | 4842 (90.30) |  |
| High school or vocational school | 1510 (9.38) | 519 (9.56) | 559 (10.53) | 432 (8.06) |  |
| Above high school or vocational school | 275 (1.71) | 77 (1.42) | 110 (2.07) | 88 (1.64) |  |
| Marital status (%) |  |  |  |  | <0.001 |
| Married | 13308 (82.65) | 4507 (83.02) | 4431 (83.45) | 4370 (81.48) |  |
| Married separation | 759 (4.72) | 254 (4.68) | 275 (5.18) | 230 (4.29) |  |
| Separated | 66 (0.41) | 31 (0.57) | 23 (0.43) | 12 (0.22) |  |
| Divorced | 135 (0.84) | 65 (1.20) | 42 (0.79) | 28 (0.52) |  |
| Widowed | 1710 (10.62) | 495 (9.12) | 511 (9.62) | 704 (13.13) |  |
| Never married | 124 (0.77) | 77 (1.42) | 28 (0.53) | 19 (0.35) |  |
| Hypertension (%) |  |  |  |  | <0.001 |
| yes | 4653 (28.94) | 937 (17.26) | 1432 (26.97) | 2284 (42.59) |  |
| no | 11449 (71.06) | 4492 (82.74) | 3878 (73.03) | 3079 (57.41) |  |
| Diabetes Mellitus (%) |  |  |  |  | <0.001 |
| yes | 1138 (7.22) | 172 (3.21) | 357 (6.84) | 609 (11.60) |  |
| no | 14694 (92.78) | 5189 (96.79) | 4865 (93.16) | 4640 (88.40) |  |
| Dyslipidemia (%) |  |  |  |  | <0.001 |
| yes | 1898 (12.17) | 316 (5.97) | 579 (11.20) | 1003 (19.35) |  |
| no | 13747 (87.83) | 4977 (94.03) | 4590 (88.80) | 4180 (80.65) |  |
| **Cardiovascular Disease (%)** |  |  |  |  | <0.001 |
| yes | 2156 (13.61) | 526 (9.79) | 665 (12.70) | 965 (18.33) |  |
| no | 13720 (86.39) | 4849 (90.21) | 4571 (87.30) | 4300 (81.67) |  |
| **Stroke (%)** |  |  |  |  | <0.001 |
| yes | 444 (2.79) | 124 (2.30) | 133 (2.54) | 187 (3.54) |  |
| no | 15469 (97.21) | 5267 (97.70) | 5109 (97.46) | 5093 (96.46) |  |
| **SBP, mmHg** | 128.46 ± 19.60 | 123.87 ± 19.23 | 127.84 ± 19.46 | 133.68 ± 20.12 | <0.001 |
| **DBP, mmHg** | 75.10 ± 11.31 | 72.55 ± 11.26 | 75.08 ± 11.30 | 77.67 ± 11.37 | <0.001 |
| **PP, mmHg** | 53.36 ± 14.25 | 51.32 ± 13.31 | 52.76 ± 13.88 | 56.01 ± 15.55 |  |
| **BMI, Kg/m2** | 23.60 ± 2.91 | 20.62 ± 2.66 | 23.36 ± 2.63 | 26.83 ± 3.45 | <0.001 |
| **Antihypertensive medication (%)** |  |  |  |  | <0.001 |
| yes | 3326 (20.92) | 601 (11.15) | 972 (18.48) | 1753 (33.12) |  |
| no | 12617 (79.08) | 4788 (88.85) | 4289 (81.52) | 3540 (66.88) |  |
| **Antidiabetic medication (%)** |  |  |  |  | <0.001 |
| yes | 761 (4.84) | 116 (2.17) | 227 (4.36) | 418 (8.00) |  |
| no | 15020 (95.16) | 5231 (97.83) | 4981 (95.64) | 4808 (92.00) |  |
| **Drinking (%)** |  |  |  |  | <0.001 |
| yes | 6949 (43.15) | 2795 (51.52) | 2347 (44.21) | 1807 (33.73) |  |
| no | 9142 (56.85) | 2630 (48.48) | 2962 (55.79) | 3550 (66.27) |  |
| **Smoking (%)** |  |  |  |  | <0.001 |
| yes | 6673 (41.40) | 2998 (55.25) | 2177 (41.00) | 1498 (27.96) |  |
| no | 9421 (58.60) | 2428 (44.75) | 3133 (59.00) | 3860 (72.04) |  |
| **Laboratory indices** |  |  |  |  |  |
| **TC, mg/dL** | 190.58 ± 38.01 | 183.73 ± 36.68 | 190.66 ± 37.47 | 197.34 ± 39.87 | <0.001 |
| **TG, mg/dL** | 135.41 ± 96.78 | 104.59 ± 66.27 | 137.00 ± 101.95 | 164.64± 122.12 | <0.001 |
| **LDL-C, mg/dL** | 112.20 ± 33.61 | 107.38 ± 32.11 | 112.26 ± 33.12 | 116.95 ± 35.61 | <0.001 |
| **HDL-C, mg/dL** | 51.21 ± 13.69 | 55.86 ± 15.04 | 50.62 ± 13.72 | 47.14 ± 12.32 | <0.001 |
| **FBG, mmol/L** | 5.99 ± 1.95 | 5.71 ± 1.60 | 5.98 ± 1.90 | 6.28 ± 2.36 | <0.001 |
| **BUN, mg/dL** | 15.66 ± 4.61 | 15.97 ± 4.75 | 15.58 ± 4.73 | 15.44 ± 4.36 | <0.001 |
| **SUA, mg/dL** | 4.62 ± 1.33 | 4.50 ± 1.26 | 4.64 ± 1.37 | 4.73 ± 1.35 | <0.001 |
| **Scr, mg/dL** | 0.79 ± 0.26 | 0.81 ± 0.22 | 0.80 ± 0.35 | 0.77 ± 0.21 | <0.001 |
| **CRP, mg/dL** | 2.75 ± 7.13 | 2.44 ± 6.90 | 2.65 ± 7.56 | 3.16 ± 6.94 | <0.001 |
| **HbA1c (%)** | 5.49 ± 0.91 | 5.31 ± 0.77 | 5.46 ± 0.89 | 5.70 ± 1.08 | <0.001 |

Annotation: Data are presented as mean ± standard deviation or number (%).

### Association Between BRI Z-score and Hypertension Prevalence

Table 2 illustrates the association between BRI Z-score and HTN prevalence. This relationship was confirmed through regression analysis using three different models. In Model 1, the odds ratio (OR) for the BRI Z-score was 1.70 (95% CI: 1.64–1.77, p < 0.0001), indicating that for every one-unit increase in the BRI Z-score, HTN prevalence increased by 70%. In Model 2 and Model 3, after stepwise adjustments for confounding factors, the ORs remained significant, at 1.70 and 1.58, respectively (p < 0.0001). In different BRI Z-score groups, the HTN prevalence in the Q2 and Q3 groups was significantly higher than in the Q1 group, with the highest risk observed in the Q3 group. In Model 1, the OR for the Q3 group was 3.56 (95% CI: 3.25–3.89), and after adjustment in Model 3, the OR remained at 3.09 (95% CI: 2.76–3.47). These results suggest that higher BRI Z-scores are associated with higher HTN prevalence, a relationship that remains significant even after adjusting for multiple confounders.

**Table 2.** Multivariate regression analysis of the association between BRI Z-score and hypertension prevalence.

|  |  | Model 1 | Model 2 | Model 3 |
| --- | --- | --- | --- | --- |
| Variable | Group | OR (95% CI), *p*-value | OR (95% CI), *p*-value | OR (95% CI), *p*-value |
| BRI Z-score | Continuous | 1.70 (1.64, 1.77), <0.0001 | 1.70 (1.64, 1.77), <0.0001 | 1.58 (1.51, 1.66), <0.0001 |
|  | Q1 | Ref. | Ref. | Ref. |
| BRI Z-score groups | Q2 | 1.77 (1.61, 1.94), <0.0001 | 1.86 (1.69, 2.04), <0.0001 | 1.65 (1.47, 1.84), <0.0001 |
|  | Q3 | 3.56 (3.25, 3.89), <0.0001 | 3.67 (3.34, 4.03), <0.0001 | 3.09 (2.76, 3.47), <0.0001 |

Model 1: no covariates were adjusted.
Model 2: Age, gender, education level, smoking status, drinking status and marital status were adjusted.
Model 3: adjusted for LDL-C, HDL-C, TG, TC on the basis of Model 2.
BRI analyzed as a continuous variable and in three groups: Q1 (BRI Z-score ≤ -0.436), Q2 (−0.436 > BRI Z-score ≤ 0.355), and Q3 (BRI Z-score > 0.355).

### Non-linear relationship between BRI Z-score and Hypertension prevalence

The results from the RCS curve and threshold effect analysis indicate a nonlinear relationship between BRI Z-score and HTN prevalence. After adjusting for all covariates, Figure 4 shows that HTN prevalence remains stable at lower BRI Z-scores but increases significantly as the BRI Z-score approaches 0. This nonlinear relationship is statistically significant (P for nonlinear < 0.001), indicating that higher BRI Z-scores are associated with an increased prevalence of HTN. In the threshold effect analysis presented in Table 3, Model 1’s linear model shows that each one-unit increase in BRI Z-score is associated with a 70% increase in HTN prevalence (OR = 1.70, p < 0.0001). Model 2 identifies a threshold at -1.3, where HTN prevalence is lower for BRI Z-scores below -1.3 (OR = 0.62, p < 0.0001) and significantly higher for BRI Z-scores above -1.3 (OR = 1.89, p < 0.0001). These findings support the presence of a nonlinear and threshold effect between BRI Z-score and HTN prevalence.

**Table 3.** Threshold effect analysis of BRI Z-score on hypertension prevalence.

| BRI Z-score | Hypertension prevalence |
| --- | --- |
| Model 1 |  |
| Fitting by the standard linear model | 1.70 (1.64, 1.77), <0.0001 |
| Model 2 |  |
| Inflection point | -1.3 |
| <-1.3 | 0.62 (0.54, 0.71), <0.0001 |
| > -1.3 | 1.89 (1.81, 1.97), <0.0001 |
| Log-likelihood ratio | <0.001 |

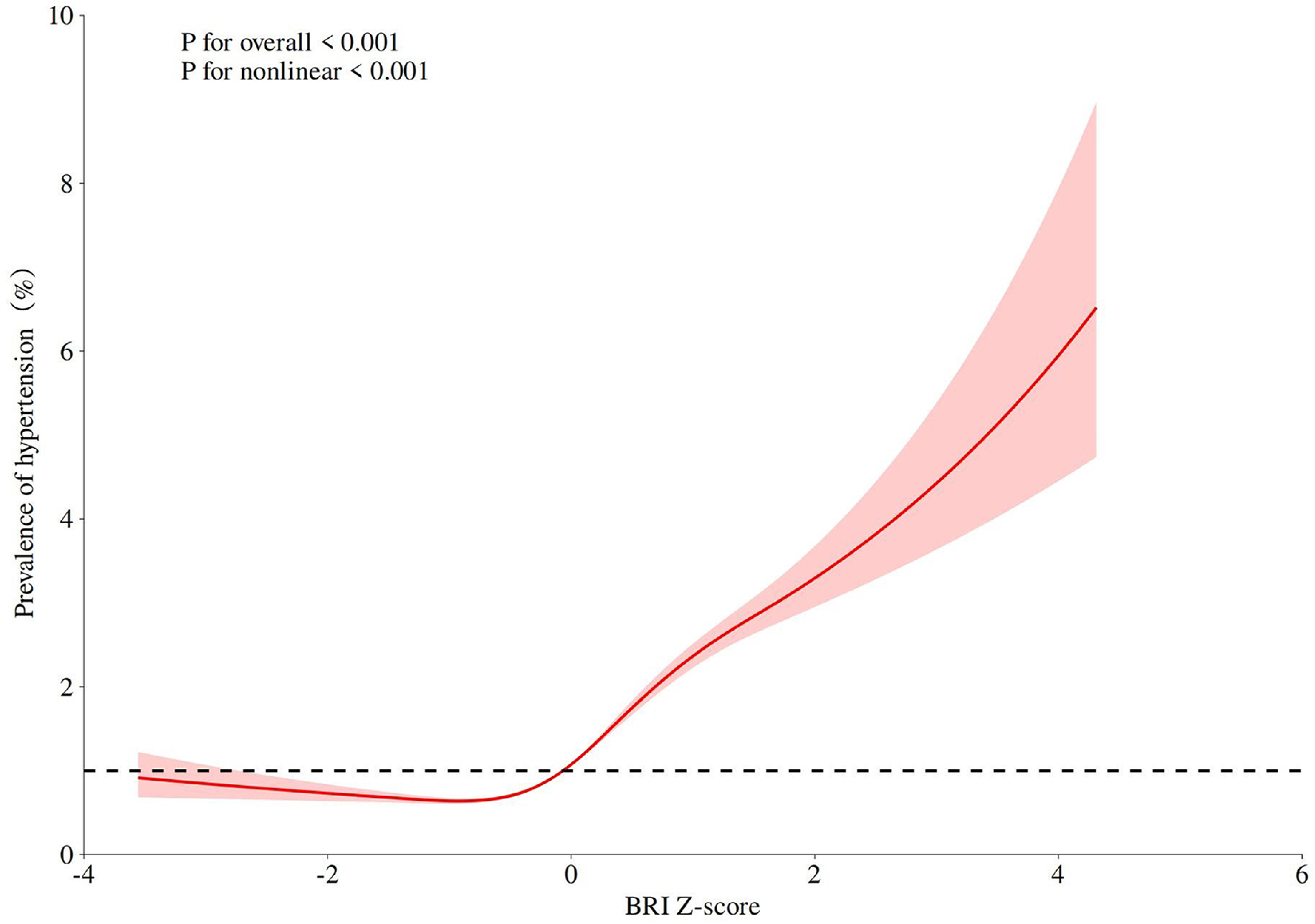

### Subgroup analysis

To ensure the reliability of the results, subgroup analyses were conducted. Figure 5 presents the associations between the BRI Z-score and HTN, SBP, DBP, and PP across different subgroups. Overall, the BRI Z-score was significantly positively correlated with all blood pressure indicators (P < 0.001). Specifically, gender, age, diabetes history, and dyslipidemia significantly influenced the association between the BRI Z-score and blood pressure. Notably, stronger associations were observed in females, individuals under 60 years of age, and those without a history of diabetes or dyslipidemia, with significant interaction effects for gender and diabetes history (P < 0.05). These results indicate that the relationship between the BRI Z-score and hypertension varies by population characteristics, highlighting the importance of personalized assessment.

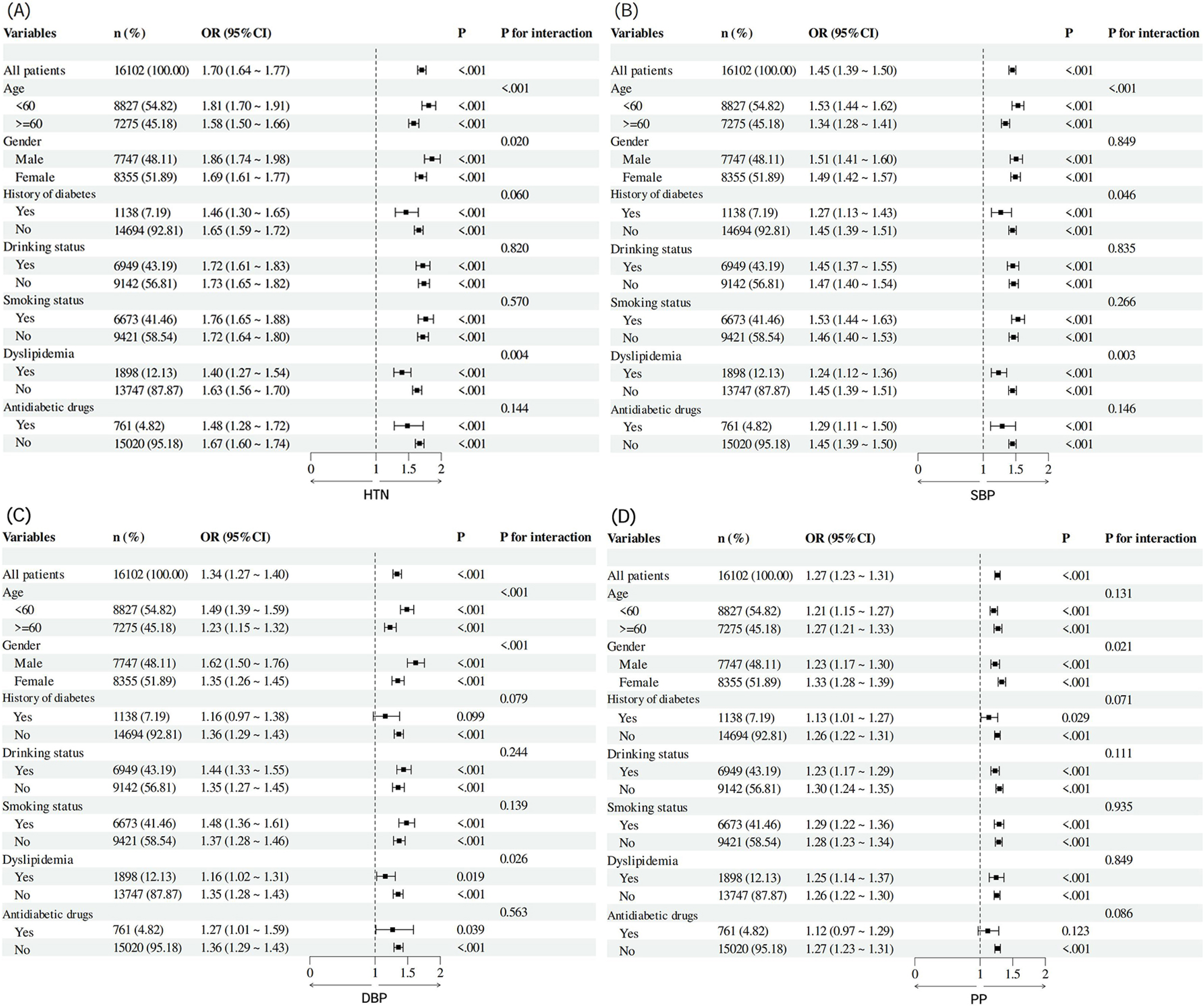

### Association Between BRI Z-score and Blood Pressure Measurement

Table 4 examines the association between BRI Z-score and blood pressure, with the HTN diagnostic threshold adjusted to 130/80 mmHg[13]. The results indicate that BRI Z-score remains a significant predictor under this revised standard. For the traditional HTN criteria (SBP ≥ 140 mmHg or DBP ≥ 90 mmHg), each one-unit increase in BRI Z-score was associated with a 42% and 47% increased risk of elevated SBP and DBP, respectively (Model 3 ORs: 1.42 and 1.47, both p < 0.0001). In subgroup analyses, the Q3 group exhibited the highest risk, with ORs of 2.31 for SBP and 2.45 for DBP (95% CI: 2.05–2.60 and 2.08–2.89, respectively). Within the new diagnostic threshold[14, 15], we observed that each one-unit increase in BRI Z-score was associated with an 12% and 29% increased risk of SBP 130–140 mmHg and DBP 80–90 mmHg, respectively (Model 3 ORs: 1.11 and 1.29, both p < 0.0001). Subgroup analyses further revealed a significant risk increase in the Q3 group, with ORs of 1.41 for SBP 130–140 mmHg and 1.89 for DBP 80–90 mmHg (95% CI: 1.23–1.61 and 1.67–2.13, respectively). This robust association suggests that BRI Z-score is not only an effective predictor of traditional HTN but also a valuable tool for identifying early blood pressure risk, particularly within the borderline-elevated range (130–140 mmHg and 80–90 mmHg). These findings underscore the importance of early intervention in individuals with high BRI Z-scores.

**Table 4.** Multivariate regression analysis of BRI Z-score status and indicators of blood pressure.

| Variable | Group | Model 1 | Model 2 | Model 3 |
| --- | --- | --- | --- | --- |
|  |  | OR (95% CI), *p*-value | OR (95% CI), *p*-value | OR (95% CI), *p*-value |
| 140 mmHg ≤ SBP | Continuous | 1.45 (1.39, 1.50), < 0.0001 | 1.45 (1.39, 1.51), < 0.0001 | 1.42 (1.35, 1.49), < 0.0001 |
|  | Q1 | Ref. | Ref. | Ref. |
|  | Q2 | 1.40 (1.27, 1.53), < 0.0001 | 1.51 (1.37, 1.67), < 0.0001 | 1.39 (1.24, 1.56), < 0.0001 |
|  | Q3 | 2.36 (2.16, 2.57), <0.0001 | 2.48(2.25, 2.73), <0.0001 | 2.31 (2.05, 2.60), <0.0001 |
| 90 mmHg ≤ DBP | Continuous | 1.34 (1.27, 1.40), < 0.0001 | 1.51 (1.43, 1.59), <0.0001 | 1.47 (1.38, 1.58), < 0.0001 |
|  | Q1 | Ref. | Ref. | Ref. |
|  | Q2 | 1.44 (1.26, 1.64), < 0.0001 | 1.58 (1.38, 1.81), <0.0001 | 1.42 (1.21, 1.68), < 0.0001 |
|  | Q3 | 2.12 (1.87, 2.41), < 0.0001 | 2.71 (2.37, 3.09), < 0.0001 | 2.45 (2.08, 2.89), < 0.0001 |
| 130 mmHg ≤ SBP < 140 mmHg | Continuous | 1.11 (1.07, 1.16), < 0.0001 | 1.15 (1.10, 1.20), < 0.0001 | 1.12 (1.06, 1.18), < 0.0001 |
|  | Q1 | Ref. | Ref. | Ref. |
|  | Q2 | 1.19 (1.07, 1.32), < 0.0001 | 1.24 (1.11, 1.38), < 0.0001 | 1.26 (1.11, 1.43), 0.0004 |
|  | Q3 | 1.34 (1.21, 1.49), <0.0001 | 1.44 (1.30, 1.61), <0.0001 | 1.41 (1.23, 1.61), <0.0001 |
| 80 mmHg ≤ DBP < 90 mmHg | Continuous | 1.22 (1.17, 1.26), < 0.0001 | 1.28 (1.23, 1.33), <0.0001 | 1.29 (1.23, 1.36), < 0.0001 |
|  | Q1 | Ref. | Ref. | Ref. |
|  | Q2 | 1.35 (1.23, 1.49), < 0.0001 | 1.41 (1.28, 1.56), <0.0001 | 1.40 (1.24, 1.57), < 0.0001 |
|  | Q3 | 1.69 (1.54, 1.86), < 0.0001 | 1.88 (1.71, 2.08), < 0.0001 | 1.89 (1.67, 2.13), < 0.0001 |

Model 1: no covariates were adjusted.
Model 2: Age, gender, education level, smoking status, drinking status and marital status were adjusted.
Model 3: adjusted for LDL-C, HDL-C, TG, TC on the basis of Model 2.
BRI analyzed as a continuous variable and in three groups: Q1 (BRI Z-score ≤ -0.436), Q2 (−0.436 > BRI Z-score ≤ 0.355), and Q3 (BRI Z-score > 0.355).

## Discussion

In recent years, BMI, a traditional measure of obesity and cardiovascular risk, has shown its limitations[16]. Although BMI is widely used for its simplicity, it fails to effectively differentiate between fat and lean tissue and does not account for fat distribution[17]. The distribution of fat, especially abdominal fat accumulation, is crucial when assessing the risks of metabolic and cardiovascular diseases[4, 18]. Consequently, indicators like waist circumference, waist-to-height ratio, and the BRI have gradually been recognized as more accurate tools for evaluating central obesity[19]. Specifically, BRI provides a comprehensive index by combining waist circumference and height, offering a more precise measure of body shape and fat distribution[20]. Furthermore, the introduction of the BRI Z-score, which standardizes values across different populations, enhances the utility of this measure, making risk assessment and comparison more precise.

HTN, a significant global public health issue, affects over a billion people and leads to high morbidity and mortality rates[21]. With the acceleration of urbanization, population aging, and lifestyle changes, HTN prevalence continues to rise, especially in low- and middle-income countries[22]. The close relationship between central obesity and HTN has become a focal point. Our study demonstrates that the BRI Z-score, which assesses central obesity, is significantly positively correlated with HTN, particularly in the middle-aged and elderly populations, highlighting the urgency of HTN prevention and management.

Obesity, especially central obesity, is widely recognized as a major risk factor for HTN[23, 24]. Unlike generalized obesity, central obesity is characterized by visceral fat accumulation, which is metabolically more active than subcutaneous fat and is closely linked to adverse cardiovascular outcomes[25, 26]. Visceral fat promotes the development of HTN through multiple mechanisms, including the activation of the renin-angiotensin-aldosterone system, chronic low-grade inflammation, insulin resistance, and excessive activation of the sympathetic nervous system[27, 28]. These mechanisms collectively contribute to vasoconstriction, fluid retention, and endothelial dysfunction, thereby facilitating the development of HTN[29]. Research emphasizes that public health interventions targeting central obesity not only reduce HTN risk but also alleviate the burden of non-communicable diseases[30].

Geographical analysis reveals significant regional differences in BRI and HTN prevalence. As shown in Figure 2, Xinjiang has the highest BRI value, indicating a more severe central obesity issue in the region, while Tibet and some western areas exhibit lower BRI values, reflecting regional disparities. Figure 3 demonstrates higher hypertension prevalence in provinces such as Henan and Sichuan, compared to lower rates in Tibet and certain western regions. These differences highlight the critical role of lifestyle, dietary habits, socioeconomic status, and healthcare accessibility in shaping regional health disparities. Consequently, targeted public health strategies are essential. In regions with high BRI values, such as Xinjiang, Henan, and Sichuan, priority should be given to improving diet and promoting physical activity. In contrast, for areas with lower BRI values, such as Tibet, maintaining healthy lifestyles is crucial to prevent the emergence of central obesity and hypertension associated with rapid urbanization and lifestyle changes.

A key finding of this study is the non-linear relationship between the BRI Z-score and HTN. Using restricted cubic spline analysis, we found that the risk of HTN remained relatively stable when the BRI Z-score was low but increased dramatically as the BRI Z-score approached 0 or higher. This finding suggests that even a slight increase in central fat, once it surpasses a certain threshold, significantly elevates blood pressure. This result is consistent with known pathophysiological mechanisms, as a higher BRI Z-score reflects greater central fat accumulation and is closely associated with elevated systolic and diastolic blood pressure levels and HTN prevalence[31]. Even moderate increases in central fat can significantly heighten HTN risk, underscoring the need for early HTN detection and intervention[32].

Additionally, the association between the BRI Z-score and HTN varies across different populations. For example, this association is more pronounced in women, younger individuals, and those without diabetes or dyslipidemia. These differences suggest that some groups may be more susceptible to the adverse effects of central obesity on blood pressure regulation. In women, hormonal changes, particularly after menopause, may exacerbate the impact of central fat on vascular function and blood pressure[33]. Estrogen has a protective effect on the cardiovascular system, and its decline after menopause may increase visceral fat-related inflammation and metabolic effects[34]. In younger populations, a higher BRI Z-score may indicate a lifetime risk of HTN and associated complications, further emphasizing the importance of early prevention.

Public health measures can utilize the BRI Z-score to identify high-risk individuals and implement appropriate interventions. For example, young women with a high BRI Z-score, even without other metabolic abnormalities, should receive lifestyle interventions to prevent HTN. The simplicity and cost-effectiveness of the BRI Z-score make it an ideal tool for large-scale screening, particularly in low- and middle-income countries (LMICs)[35]. Community health workers can easily measure waist circumference and height, calculate the BRI Z-score, and identify high-risk individuals. Incorporating the BRI Z-score into existing public health programs, such as obesity prevention or cardiovascular risk assessment, can enhance the effectiveness of health interventions and promote early HTN detection.

This study also emphasizes the relationship between the BRI Z-score and pre-HTN (SBP 130–140 mmHg or DBP 80–90 mmHg). Elevated BRI Z-scores were associated with an increased prevalence of pre-HTN, suggesting that central obesity may induce early vascular changes before overt HTN manifests. This finding highlights the importance of early detection and management of central obesity in individuals with blood pressure approaching critical levels[36]. Lifestyle interventions, particularly measures to reduce central fat, can prevent the progression from pre-HTN to overt HTN, thereby reducing the long-term cardiovascular disease burden.

Although this study provides valuable insights into the relationship between the BRI Z-score and hypertension, several limitations should be noted. First, cross-sectional design limits causal inference, and longitudinal studies are needed to confirm its predictive role. Second, regional diversity in the sample may affect the generalizability of the results, and further validation in different populations is necessary. Third, variations in hypertension definitions and measurement errors may influence the findings, and future research should standardize blood pressure measurement while considering additional confounding factors. Finally, the BRI Z-score calculation relies on waist circumference and height, and its applicability in different populations should be optimized in future studies.

## Conclusion

In conclusion, this study highlights the limitations of BMI in assessing the relationship between body fat distribution and HTN risk and underscores the potential of the BRI Z-score as a promising alternative. The BRI Z-score not only enhances the precision of individual risk assessments but also offers valuable insights for optimizing public health strategies. With the growing global burden of obesity and its related health complications, incorporating the BRI Z-score into preventive health strategies may be a crucial step toward more effective HTN management and improved cardiovascular health outcomes.

### Perspectives

This study underscores the critical role of central obesity, measured by the BRI Z-score, in predicting HTN risk, providing a fresh perspective on obesity-related health assessments. Unlike BMI, the BRI Z-score accurately captures fat distribution and its association with HTN, addressing a long-standing gap in traditional metrics. The non-linear relationship observed between BRI Z-score and HTN prevalence highlights the significant impact of even moderate increases in central fat on blood pressure, emphasizing the importance of early detection and intervention. The findings have far-reaching implications for public health and clinical practice, offering a simple, cost-effective tool for large-scale screenings and targeted interventions, particularly in resource-limited settings. Subgroup analysis reveals that high-risk populations, such as younger individuals, women, and those without metabolic disorders, may benefit most from personalized strategies based on BRI Z-scores. Integrating the BRI Z-score into existing HTN prevention programs can facilitate timely lifestyle interventions, reduce cardiovascular risk, and address the global burden of obesity-related complications.

### Clinical Perspective

1. What Is New? The BRI Z-score is a more precise measure of central obesity than BMI, showing a strong, non-linear association with hypertension risk. It effectively identifies early blood pressure abnormalities (pre-hypertension), enabling earlier intervention.
2. What Are the Clinical Implications? The BRI Z-score improves hypertension risk prediction, particularly in those with central obesity, supports early detection of pre-hypertension for timely interventions, and offers a simple, cost-effective tool for large-scale public health screenings.

## Acknowledgements

We would like to thank the initiators and participants of the CHARLS database.

## Author contributions

**Liangxiu Wu, Shenshen Du, Weicheng Lai**: Writing – review & editing, Writing − original draft, Validation, Supervision, Project administration, Methodology. **Mengxuan Liu, Yupeng Wu, Huayang Qin, Pinyou Lu, Qimeng Wu, Qian Liu, Qiao Jie**: Writing − review & editing, Validation, Investigation, Formal analysis. **Xin Li, Liangyan Wu:** Writing − review & editing, Writing − original draft, Resources, Project administration, Formal analysis, Data curation, Conceptualization.

## Funding

This work was supported by the Elderly Health Research Project from the Jiangsu Provincial Health Commission (LKM2023029).

## Data availability

The CHARLS data that support the findings of this study are openly available at https://charls.pku.edu.cn/.

## Declarations

### Ethics approval and consent to participate

The CHARLS protocol was approved by the Biomedical Ethics Review Committee of Peking University (IRB00001052-11014) and the Institutional Review Board of the National School of Development at Peking University (IRB00001052-11015), with informed consent obtained from all participants.

### Consent for publication

Not applicable.

### Competing interests

The authors declare no competing interests.

## Notes

### Competing Interest Statement

The authors have declared no competing interest.

